# A pro-oxidant combination of resveratrol and copper down-regulates hallmarks of cancer and immune checkpoints in patients with advanced oral cancer: Results of an exploratory study (RESCU 004)

**DOI:** 10.1101/2022.07.21.22277851

**Authors:** Aishwarya Pilankar, Hitesh Singhavi, Gorantla V. Raghuram, Sophiya Siddiqui, Naveen Kumar Khare, Vishalkumar Jadhav, Harshali Tandel, Kavita Pal, Atanu Bhattacharjee, Pankaj Chaturvedi, Indraneel Mittra

## Abstract

**Background:** Our earlier studies have shown that cell-free chromatin particles (cfChPs) that are released from dying cancer cells are readily internalised by bystander cells leading to activation of two hallmarks of cancer viz. genome instability and inflammation. These hallmarks could be down-regulated by deactivating cfChPs via medium of oxygen radicals generated upon admixing small quantities of the nutraceuticals resveratrol (R) and copper (Cu). In this exploratory study, we investigated whether oral administration of R and Cu (R-Cu) would down-regulate the hallmarks of cancer and immune checkpoints in advanced squamous cell carcinoma of oral cavity (OSCC).

**Patients and methods:** The study comprised of 25 patients divided into 5 equal groups. Five patients acted as controls; the remaining 20 were given R-Cu in four escalating doses. The lowest dose of R-Cu was 5.6mg and 560ng respectively, and the highest dose was 500mg and 5mg respectively. An initial biopsy was taken from patients at first presentation, and a second biopsy was taken 2 weeks later on the operating table. R-Cu was administered orally twice daily in the intervening period. Confocal microscopy was performed on tumour sections after fluorescent immuno-staining with anti-DNA and anti-histone antibodies to detect presence of cfChPs in the tumour micro-environment (TME). Immunofluorescence analysis was performed for 23 biomarkers representing the 10 Hallmarks of cancer, including 5 immune checkpoints, defined by Hanahan and Weinberg.

**Results:** Confocal microscopy detected copious presence of cfChPs in TME of OSCC, which were eradicated / deactivated following two-week treatment with R-Cu. Eradication of cfChPs from TME was associated with marked down-regulation of 21 / 23 biomarkers, including the five immune checkpoints. The lower two doses of R-Cu were more effective than the higher doses. No adverse effects attributable to R-Cu were observed.

**Conclusion:** These results suggest that cfChPs released into TME from dying cancer cells are global instigators for cancer hallmarks and immune checkpoints in surviving cancer cells. The ability of R-Cu to deactivate cfChPs raises the prospect of a novel and non-toxic form of cancer treatment which sans killing of cancer cells, and instead induces healing by down-regulating cancer hallmarks and immune check-points.

**Clinical Trial Registration:** http://ctri.nic.in/Clinicaltrials/login.php,(CTRI/2018/03/012459)

## Introduction

Results of treatment of advanced squamous cell carcinoma of oral cavity (OSCC) continue to remain unsatisfactory, and is associated with considerable toxic side effects. Novel therapeutic approaches that are non-toxic are urgently needed. Our earlier studies have shown that cell-free chromatin particles (cfChPs) that are released from dying cancer cells can readily enter into surrounding bystander cells leading to activation of two critical hallmarks of cancer viz. dsDNA breaks (genome instability) and inflammation, which could lead to their oncogenic transformation (1). Activation of these cancer hallmarks could be abrogated by concurrent treatment with a combination of the nutraceuticals resveratrol (R) and copper (Cu) (1, 2). Fukuhara *et*.*al* were the first to demonstrate that oxygen radicals are generated upon admixing R and Cu (3). They showed that R acts as a catalyst to reduce Cu (II) to Cu (I) resulting in generation of oxygen radicals that are capable of cleaving plasmid pBR322 DNA (3, 4). We have extended these findings to show that oxygen radicals that are generated following admixture of R and Cu (R-Cu) can degrade genomic DNA and RNA (5), and can deactivate extra-cellular cfChPs by degrading their DNA component, both *in vitro* and *in vivo* (1, 2, 6 - 10).

Oxygen radicals are short lived molecular entities which contain an unpaired electron making them exceptionally reactive as they search for another electron to pair with, and in the process they can damage biomolecules such as DNA, proteins and lipids (11). Oxygen radical are normally generated by mitochondria (12); their over-production leads to oxidative stress which has several harmful effects on host cells (13). However, paradoxically, when oxygen radicals are artificially generated in the extracellular compartments of the body, such as by R-Cu, they can have wide ranging therapeutic effects in animal models and in human conditions that are associated with elevated levels of extracellular cfChPs (2, 6 - 10). These effects are mediated via the ability of oxygen radicals to deactivate extracellular cfChPs.

Oral administration of R-Cu leads to generation of oxygen radicals in the stomach, which are absorbed and have systemic effects in the form of deactivation of extracellular cfChPs. Our pre-clinical studies have shown that oral administration of small quantities of R-Cu can have remarkable therapeutic effects in conditions associated with elevated levels of cfChPs (2, 6-8). For example, R-Cu administered orally can prevent toxic side effects of chemotherapy (6), and radiotherapy (2), and prevent fatality in mice following bacterial endotoxin induced sepsis (7). We have also shown that prolonged oral administration of R-Cu to ageing mice can down-regulate several biological hallmarks of ageing and neurodegeneration (8). Our early results in patients receiving high dose chemotherapy and bone marrow transplantation for multiple myeloma have shown that orally administered R-Cu can significantly reduce Grade III-IV mucositis (9). Blood levels of inflammatory cytokines were also found to be significantly reduced following R-Cu treatment in that study (9). We have also shown in an observational study that oral administration of R-Cu to patients with severe Covid-19 reduced their mortality by nearly 50% (10). Significantly, we have discovered that cfChPs deactivating activity of R-Cu is retained even when the molar concentration of Cu is reduced 10,000 fold with respect to that of R (2, 5 - 10). Consequently, in our pre-clinical and clinical studies, the molar ratio of R to Cu was maintained at 1:10^−4^ (2, 6 - 10).

In the present exploratory study, we investigated whether oral administration of R-Cu would lead to down-regulation of hallmarks of cancer and immune checkpoints in patients with OSCC. We show that cfChPs that were released from dying cancer cells are abundantly present in the tumour microenvironment (TME), and that orally administered R-Cu dramatically reduced cfChPs in TME. Elimination of cfChPs from TME was associated with significant down-regulation of 21/23 biomarkers representing the 10 hallmarks of cancer, including 5 immune check-points, that have been defined by Hanahan and Weinberg. These exploratory findings suggest that prolonged treatment with R-Cu may have the potential to induce healing without having to kill cancer cells.

## Patients and Methods

### Ethics approval

This study was approved by Institutional Ethics Committee (IEC) of Advanced Centre for Treatment, Research and Education in Cancer, Tata Memorial Centre. A written informed consent was obtained from all study participants as stipulated by IEC. The trial was registered under Clinical Trial Registry of India (no.CTRI/2018/03/012459)

### Patients and R-Cu treatment

The study comprised of 25 patients with advanced OSCC who were divided into 5 groups of 5 patients each. A representative image of OSCC and H&E section of the tumour is given in Supplementary Fig.1. The first 5 patients acted as controls; the remaining 20 were given R-Cu orally, twice daily, in four escalating doses (Supplementary Table 1). Resveratrol (trade name TRANSMAX^™^) was procured from Biotivia, USA [https://www.biotivia.com/product/transmax/]; Chelated Copper was procured from Calrson Laboratories, USA [https://carlsonlabs.com/chelated-copper/]. The lowest dose of R-Cu (Dose level I) comprised of 5.6mg of R and 560ng of Cu (molar ratio 1:10^−4^). This dose was arrived at by direct conversion of dose of R and Cu that we have used in our pre-clinical studies (R = 1mg/Kg and that of Cu 0.1µg/Kg) using a standard conversion formula (14). Dose levels II and III were approximately 10 fold and 100 fold higher than dose level I; dose level IV comprised of doses of R and Cu that have been recommended by the respective vendors for use as health supplements.

R is supplied by the vendor as capsules containing 500mg of trans-resveratrol powder, and Cu is supplied as 5 mg tablets. For the purposes of this study, R powder was removed from the capsules and weighed as per requirement. Cu tablets were crushed in a pestle and mortar and the powder was weighed as per requirement. R powder being insoluble in water, and unpalatable at higher doses, was administered using honey as a vehicle (∼15 mL). Cu, being soluble in water was administered as water based solution (20mL). R and Cu were administered orally, one after the other, on empty stomach, twice daily for 2 weeks. The control group received vehicles (honey and water) alone.

A diagnostic punch biopsy was taken from patients under local anaesthesia at the time of their first presentation to Tata Memorial Hospital, and a second biopsy was taken 2 weeks later at the time of surgery on the operating table under anaesthesia. In the intervening two weeks, R-Cu was administered orally in four escalating doses as described above. The control group received vehicles alone (∼15mL honey followed by 20 mL of water). No adverse side effects attributable to R-Cu were recorded.

### Fluorescence immune-staining and confocal microscopy

Formalin fixed paraffin embedded (FFPE) sections of tumour tissues were stained with fluorescent anti-DNA and anti-histone antibodies and examined by confocal microscopy, as described by us earlier (7). This was done to investigate the presence of cfChPs (if any) in TME. Fluorescence intensity was quantified by randomly choosing five confocal fields (∼ 50 cells per field), and calculating mean fluorescence intensity (MFI) which was expressed as mean ± S.E.M.

### Immunofluorescence

Indirect immunofluorescence (IF) on FFPE sections of tumour tissues were performed according to the method described by us earlier (1). All IF analyses were performed in a blinded fashion such that the examiner was unaware of the identity of the slides being analysed. IF was used to assess the expression of 3 anti-oxidant enzymes viz. superoxide dismutase, catalase and glutathione peroxidase. Also examined by IF were 23 biomarkers representing 10 hallmarks of cancer, including 5 immune checkpoints (Supplementary Table 2). Source and catalogue numbers of antibodies used are given in (Supplementary Table 3 a - c). One thousand cells were analysed on each slide for all IF studies, and the number of positive cells was recorded. Results were expressed as mean number of positive cells (± SEM) / 1000 cells.

### Statistical analysis

Statistical comparison of pre versus post treatment values was performed in two ways: 1) combined comparison of all four treatment groups with the control group was performed applying two-way ANOVA using R software; 2) comparison of pre versus post treatment values within each treatment group was performed by paired Student’s t-test using Graphpad Prism 6.0 software.

## Results

### R-Cu treatment up-regulates anti-oxidant enzymes in tumour cells

As an initial step, we examined whether oxygen radicals generated following oral administration of R-Cu might have diffused into the tumour tissue? This was indirectly examined by IF analysis of the three antioxidant enzymes mentioned above. We observed that R-Cu treatment had led to increased expression of all three enzymes within tumour cells (Fig.1 upper panels). This indicated that oxygen-radicals had entered into the tumour cells, and that the latter in an attempt to detoxify the offending oxygen-radicals, had up-regulated anti-oxidant enzymes as a cellular defence mechanism. Combined statistical comparison of pre versus post treatment values of the four treatment groups with the control group using two-way ANOVA showed that R-Cu treatment had led to a highly significant up-regulation of all three antioxidant enzymes (Fig 1 lower panels). Individual comparison of pre and post treatment values within each treatment group using paired t test also showed significant increase in post R-Cu values for all three enzymes with respect to dose levels I – III. However, no significant difference between pre and post treatment values was seen in case of level IV for any of the three enzymes. No difference in antioxidant enzyme levels was seen between pre and post samples in the control group (Fig.1 lower panels).

**Figure 1.**
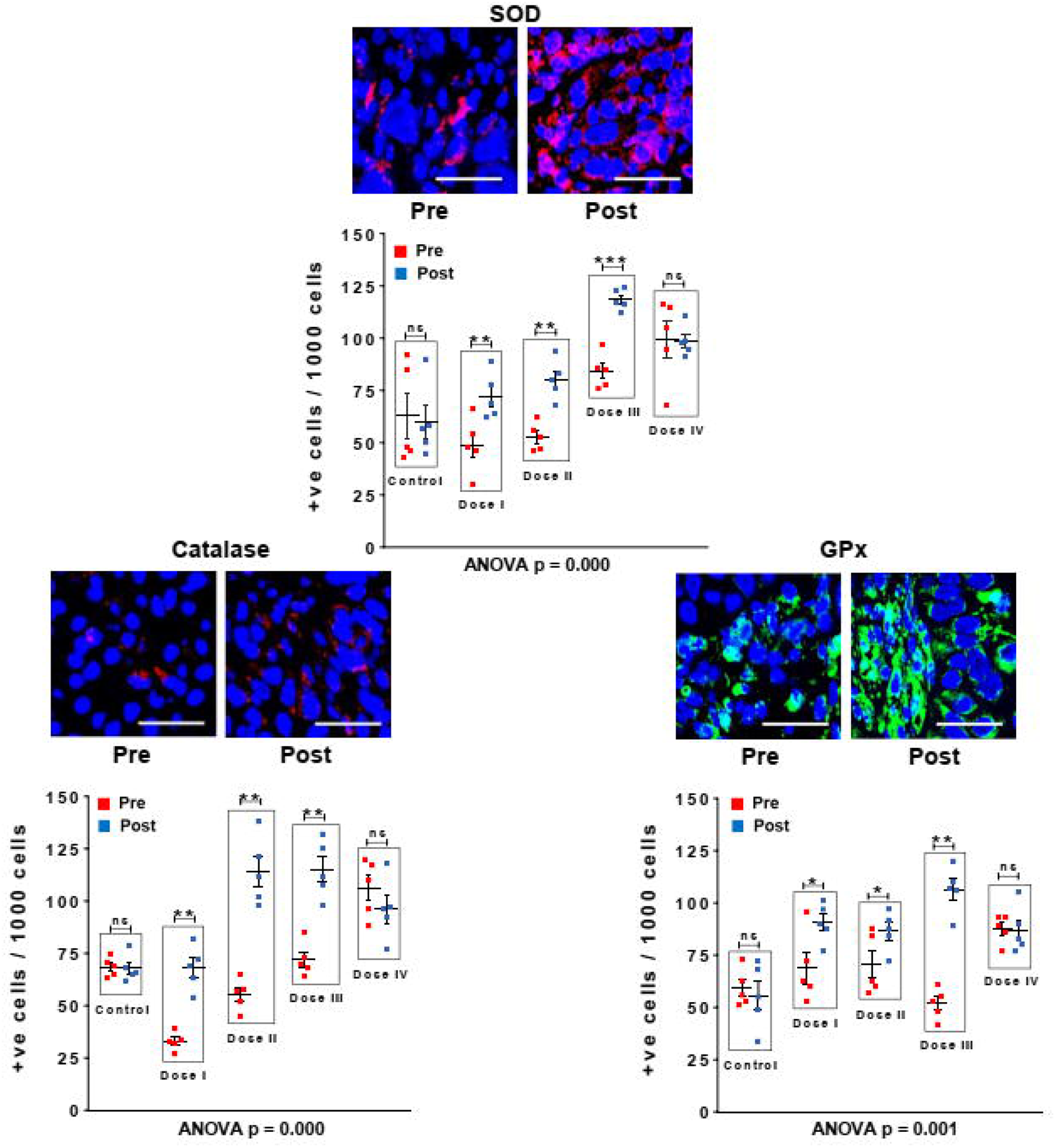
R-Cu treatment up-regulates anti-oxidant enzymes in OSCC tumour cells. FFPE sections of tumour tissues were stained with antibodies against superoxide dismutase (SOD), catalase and glutathione peroxidase (GPx), and examined under fluorescence microscopy. Upper panels provide representative images (scale bar 10µm); lower panels represent quantitative analysis of cells positive for respective anti-oxidant enzymes. One thousand cells were analysed in each case and the number of cells showing positive signals were recorded. Combined statistical comparison of pre versus post treatment values of all four treatment groups with the control group was done using two-way ANOVA; p values are given under each graph. Comparison of pre versus post treatment values within each treatment group was performed by paired Student’s t-test. Results are represented as mean ± SEM values of 5 animals in each group. * < p < 0.05; ** < p < 0.01; ***< p < 0.001; ns= not significant).

### R-Cu treatment eradicates / deactivates profusion of cfChPs present in TME

Confocal microscopy of FFPE sections of tumour tissues was performed after fluorescent immuno-staining with anti-DNA (red) and anti-histone (green) antibodies. Upon superimposing the fluorescent DNA and histone images, copious presence of yellow fluorescent signals, representing cfChPs, were detected in TME (Fig. 2, upper panel). The yellow cfChPs signals were virtually eliminated following two-week treatment with R-Cu. Mean fluorescent intensity (MFI) of yellow fluorescent cfChPs signals was quantitatively estimated and statistical analysis of the results was performed. Combined statistical comparison of pre versus post treatment MFI values of the four treatment groups with the control group using two-way ANOVA showed that treatment with R-Cu had led to a highly significant reduction in cfChPs in TME (Fig. 2, lower panel). A significant reduction in post treatment values was also seen within each treatment group when analysed by paired t test. No difference in pre and post treatment MFI values was seen in the control group (Fig. 2, lower panel). It should be noted that some of the red (DNA) and green (histone) fluorescent signals had not strictly co-localised. This is likely to be due to unevenness of cut surfaces of FFPE sections preventing the respective antibodies to have access to some of the DNA and Histone epitopes on cfChPs.

**Figure 2.**
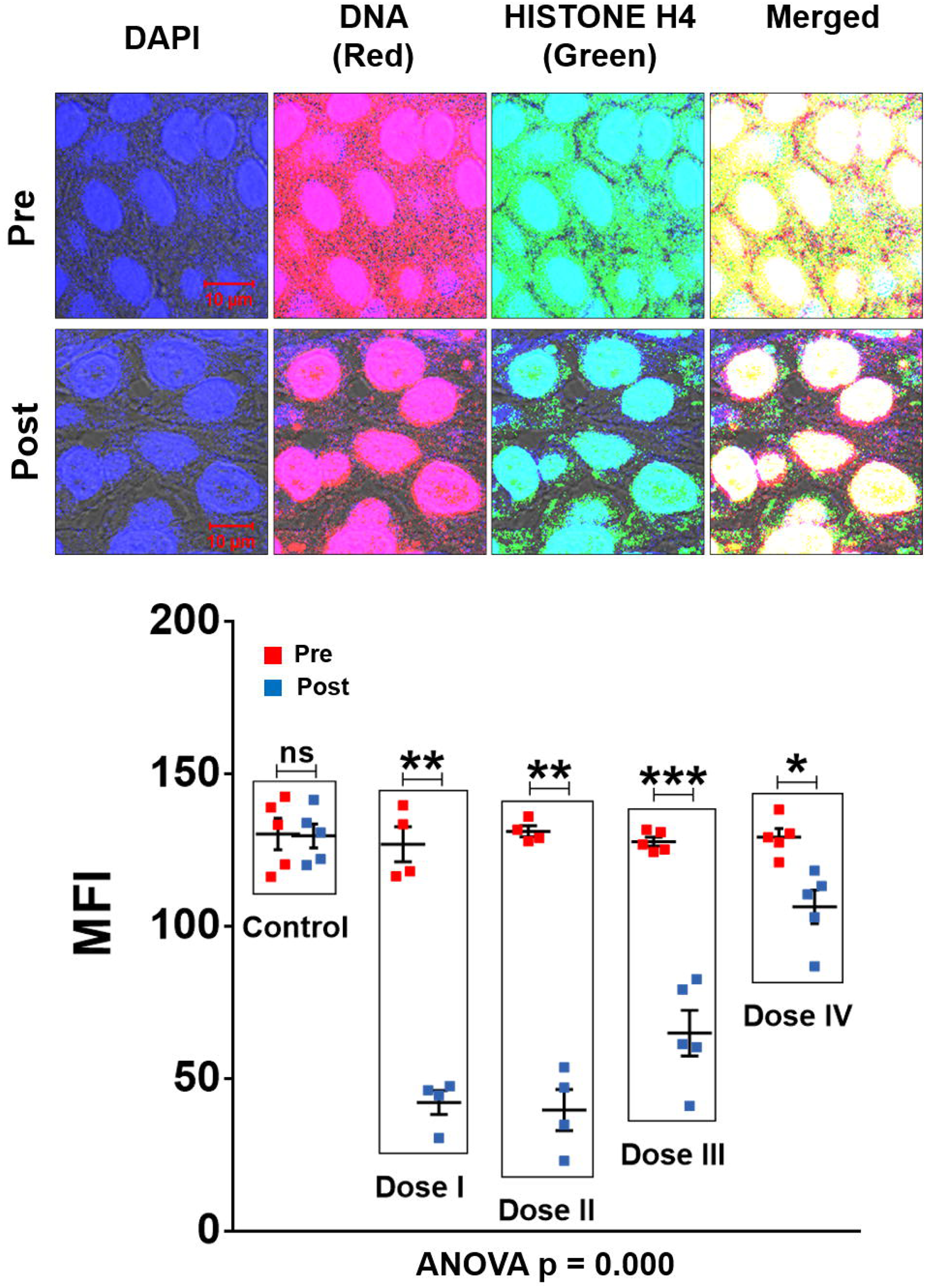
cfChPs are copiously present in TME of OSCC and are eradicated following treatment with R-Cu. Representative confocal images of FFPE sections of tumour tissues immuno-stained with anti-DNA (red) and anti-histone (green) antibodies (upper panel). Upon superimposing red and green fluorescence images, a profusion of cfCh particles (yellow) is seen in TME resulting from co-localisation of red (DNA) and green (histone) signals. Yellow cfCh signals were markedly reduced following R-Cu treatment. For quantitative analysis, MFI of five randomly chosen confocal fields (∼50 cells per field) was recorded (lower panel). Combined statistical comparison of pre versus post treatment values of all four treatment groups with the control group was done using two-way ANOVA; p values are given under the graph. Comparison of pre versus post treatment values within each treatment group was performed by paired Student’s t-test. Results are represented as mean ± SEM values of tissue sections of 5 animals in each group, except for pre and post values of dose levels I and II, wherein n was =4. * < p < 0.05; **< p < 0.01, ***< p < 0.001; ns= not significant).

### R-Cu treatment down-regulates cancer hallmarks and immune checkpoints

Comparative results of IF analysis of pre and post R-Cu treatment values of the 10 hallmarks of cancer represented by 23 biomarkers, including 5 immune checkpoints, is given in Fig.3 (upper and lower panels). Combined statistical comparison of the four treatment groups with the control group using two-way ANOVA showed a highly significant reduction in biomarker expression in the post R-Cu treatment samples in 21 / 23 cases (Fig. 3 lower panels). The exceptions were VEGFA (angiogenesis) and Glut1 (cellular energetics), wherein difference between pre and post values did not reach statistical significance. We conclude from these exploratory results that eradication of cfChPs from TME following R-Cu treatment is associated with down-regulation of 21/23 biomarkers representing 10 cancer hallmarks, and 5 immune checkpoints, that we examined.

**Figure 3.**
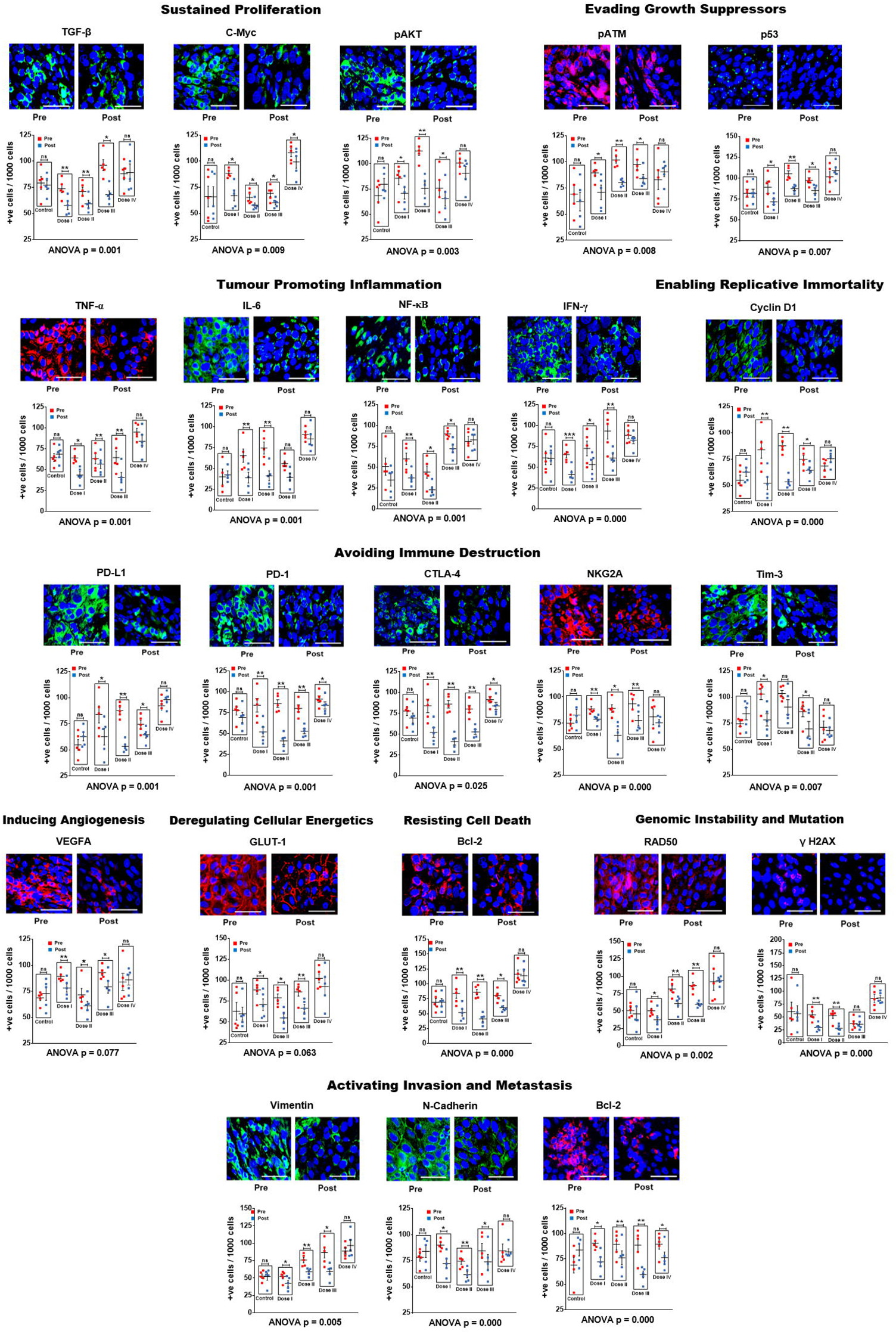
R-Cu treatment down-regulates cancer hallmark biomarkers, including immune check-points, in OSCC. Upper panels are representative IF images (scale bar 10µm); lower panels represent results of quantitative analysis of various biomarkers. One thousand cells were analysed in each case and the number of cells showing positive fluorescent signals were recorded. Combined statistical comparison of pre versus post treatment values of all four treatment groups with the control group was done using two-way ANOVA; p values are given under each graph. Comparison of pre versus post treatment values within each treatment group was performed by paired Student’s t-test. Results are represented as mean ± SEM values of tissue sections of 5 animals in each group. * < p < 0.05; ** < p < 0.01; ***< p < 0.001, ****< p < 0.0001).

Comparison of pre versus post treatment values within each treatment group using paired t-test showed that, on the whole, the two lower dose levels (I and II) were more effective. In these two treatment groups, a statistically significant reduction in biomarker expression was seen in the post treatment samples in all 23 biomarkers examined (23/23) (Fig. 3 lower panels). In case of dose level III, 20/23 and biomarkers showed statistical significant reduction in the post treatment samples. Dose level IV was least effective, with only 3/23 biomarkers showing significantly reduced values in the post R-Cu samples (Fig. 3 lower panels).

### Immune checkpoint proteins are expressed by lymphocytes present in TME

We were curious to find out whether the immune checkpoints PD-1, CTLA-4, Tim3 and NKG2A were being expressed by tumour infiltrating lymphocytes (TIL) present in TME. We performed dual IF analyses on tumour sections in which each checkpoint protein was simultaneously immune-stained for CD4 and CD8 lymphocytes (Supplementary Fig. 2 a - d). We found that all four immune checkpoint proteins co-localized with both types of immune lymphocytes. This finding confirmed that cfChPs released from the dying tumour cells had induced TIL present in TME to activate immune checkpoint proteins.

## Discussion

Results of this exploratory study need to be confirmed in a larger series of patients, and in other tumour types. Nonetheless, several observations that we made may be worthy of note. For example, that 1) oxygen radicals generated following oral administration of R-Cu were ostensibly absorbed from the stomach to enter into systemic circulation and reach the tumour cells; 2) this led the latter to up-regulate anti-oxidant enzymes as a cellular defence mechanism; 3) cfChPs are copiously present in TME having been derived from dying tumour cells; 4) oxygen radicals that diffuse into the extracellular spaces of the tumour eradicate / deactivate the profusion of cfChPs present in TME; 5) eradication of cfChPs from TME is associated with down-regulation of 21 / 23 biomarkers representing the 10 hallmarks of cancer, including 5 immune checkpoints, that have been defined by Hanahan and Weinberg (15). The mechanistic steps involved in oxygen radical induced down-regulation of cancer hallmarks are graphically illustrated in Fig.4.

**Figure 4.**
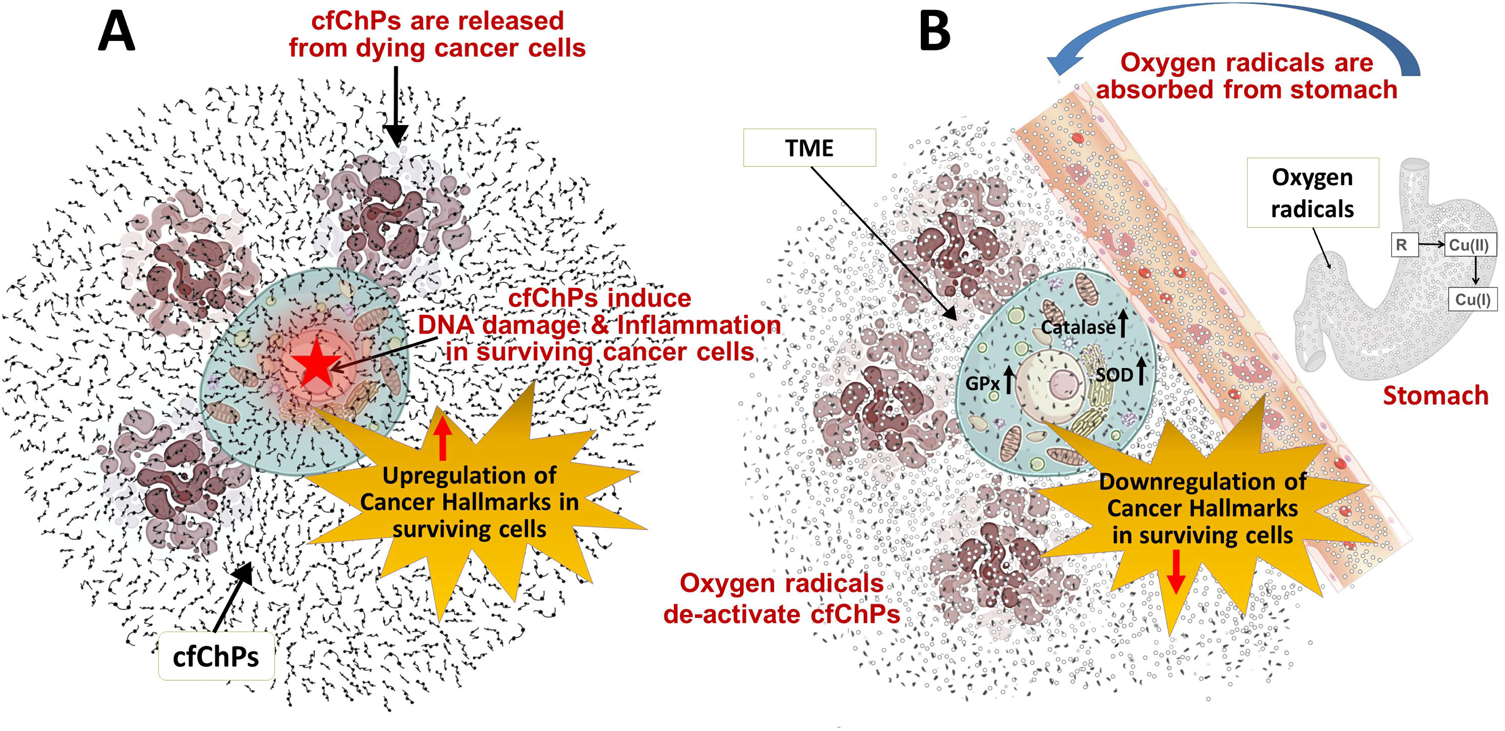
Graphical illustration of the mechanistic steps involved in oxygen radical induced down-regulation of cancer hallmarks. **A**. cfChPs released from dying cancer cells induce DNA damage and inflammation, and up-regulate cancer hallmarks, in surviving cancer cells. **B**. Oxygen radicals generated upon oral ingestion of R-Cu are systemically absorbed from the stomach to reach TME leading to eradication of extra-cellular cfChPs. Oxygen radicals also enter into the surviving cancer cells; but their cellular entry leads to activation of anti-oxidant enzymes which detoxify and eliminate the offending agents. SOD = superoxide dismutase; GPx = glutathione peroxidase; TME = tumour microenvironment.

In spite of the fact that R-Cu generated oxygen radicals had diffused into the tumour cells, we did not observe any gross damage to the tumour cell DNA following R-Cu treatment (Fig. 2 a). This suggested that the surviving tumour cells had protected themselves from damage by oxygen radical by up-regulating cellular anti-oxidant enzymes. No systemic toxic side effects were reported following R-Cu treatment for any of the dose levels administered; the up-regulated anti-oxidant enzymes had apparently protected all cells of the body from oxidative damage in a similar manner.

Results of our study suggest that cfChPs from dying cancer cells released into TME aggravate the oncogenic constitution of surviving cancer cells by activating cancer hallmarks in the tumour cells, and by activating immune checkpoint proteins in TIL. TME is known to play a critical role in all stages of tumour progression and metastases (16). It has been proposed that therapeutic targeting of constituents of TME could be a promising approach to controlling cancer (17). Our exploratory results presented herein suggest that treatment with R-Cu may be one such therapeutic approach which targets cfChPs present in TME via medium of oxygen radicals, and eradicates them to have potential therapeutic effects.

Our previous results had showed, and which are confirmed in the current study, that cellular uptake of cfChPs leads to activation of two critical hallmarks of cancer viz. dsDNA breaks (genomic instability) and inflammation (1). These two hallmarks are also crucial sensors of cellular stress triggering widespread physiological and cellular responses, which also include cancer hallmarks (18, 19). This leads us to hypothesize that cfChPs from dying cancer cells trigger a stress response in surviving cells leading to global activation of cancer hallmarks and immune checkpoints.

We observed that the two lower dose levels of R-Cu were more effective than the higher ones. It is noteworthy that the amount of R present in the lowest dose (level I) was ∼90 times less, and that of Cu ∼4500 times less, than those that are recommended by the respective vendors for use as health supplements. Thus, small quantities of a combination of R and Cu can generate sufficient oxygen radicals to have profound effects in terms of down-regulation of cancer hallmarks and immune checkpoints by targeting a hitherto unknown constituent of TME.

Our analysis showed that R-Cu treatment led to down-regulation of all five immune check-points viz. PDL-1, PD-1, CTLA-4, TIM-3, and NKG2A that were examined. Targeted therapy of immune checkpoints has drawn much recent attention, and is beginning to show promising results in cancer treatment (20). However, what triggers activation of immune checkpoints has remained known. Our exploratory results suggest that cfChPs released from dying cancer cells into TME may be global activators of immune checkpoint proteins, and that they can be down-regulated by two-week treatment with small quantities of orally administered R-Cu. If the role of cfChPs in checkpoint activation is further confirmed, a new form of cancer immunotherapy might emerge whereby all known immune checkpoints are simultaneously down-regulated by small quantities of R-Cu.

## Conclusion and future prospects

Our results suggest that cfChPs released from dying cancer cells are global instigators of cancer hallmarks and immune checkpoints. That cfChPs can be deactivated by small quantities of R-Cu raise the prospect of a novel and non-toxic form of cancer treatment which sans killing of cancer cells, and instead, induces healing by down-regulating hallmarks of cancer and immune checkpoint proteins. It has been argued that cancer is akin to a non-healing wound (21), and that hallmarks of cancer are also the hallmarks of wound healing (22). This being so, it is possible that cancer may be healed, like a wound, without having to be killed, and that oxygen radicals generated upon oral administration of R-Cu may prove to be a novel cancer healing agent of the future.

## Data Availability

All data produced in the present work are contained in the manuscript.

http://ctri.nic.in/Clinicaltrials/showallp.php?mid1=19801&EncHid=&userName=CTRI/2018/03/012459

## Acknowledgments

The authors sincerely thank Dr. Snehal Shabrish for preparing the infographic. Thanks are also due to Mr. Ashish Pawar and Mr. Roshan Shaikh for their help in preparing the manuscript.

## Author Contributions

AP: Investigation and visualization. HS: Investigation, methodology, project administration and supervision. GVR: Data curation, formal analysis, investigation, software, supervision, validation, visualization and writing–original draft. SS: Investigation and visualization. NKK: Investigation, methodology, project administration and supervision. VJ: Investigation and methodology. HT: Investigation. KP: Data curation, formal analysis, methodology, project administration, software, supervision, validation and writing–original draft. AB: Formal analysis. PC: Methodology, project administration and supervision. IM: Conceptualization, funding acquisition, methodology, project administration, resources, overall supervision, writing–original draft, writing–review and editing.

## Data Availability

All data produced in the present work are contained in the manuscript.

## Legends to supplementary figures

**Supplementary Figure 1.**
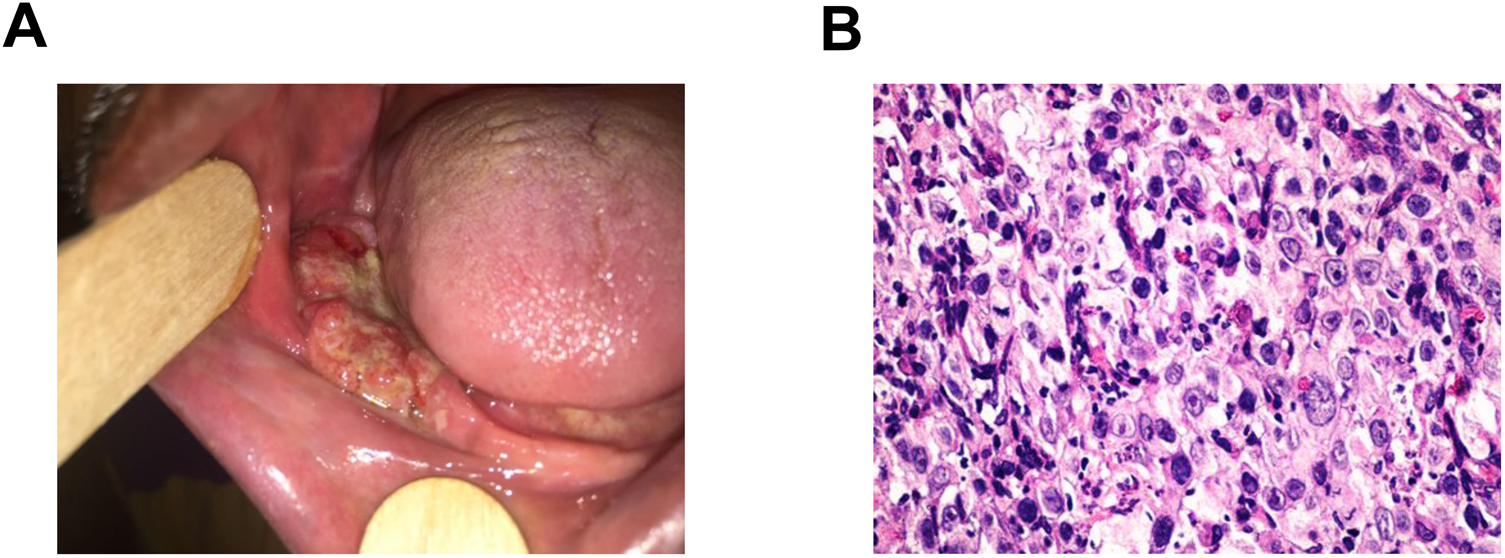
A representative photograph of advanced OSCC **A**, and its squamous cell origin as seen on a H&E section **B**.

**Supplementary Figure 2.**
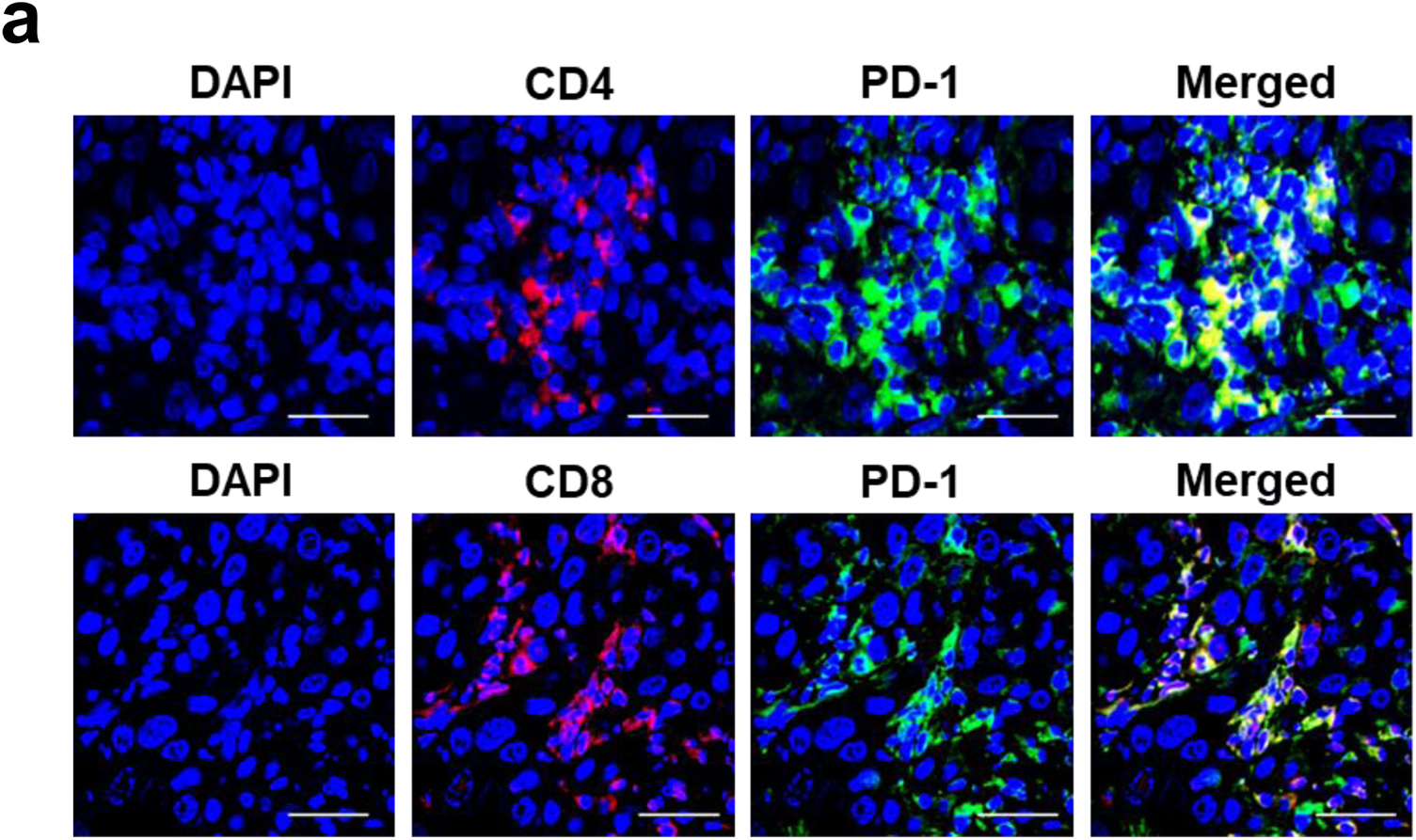
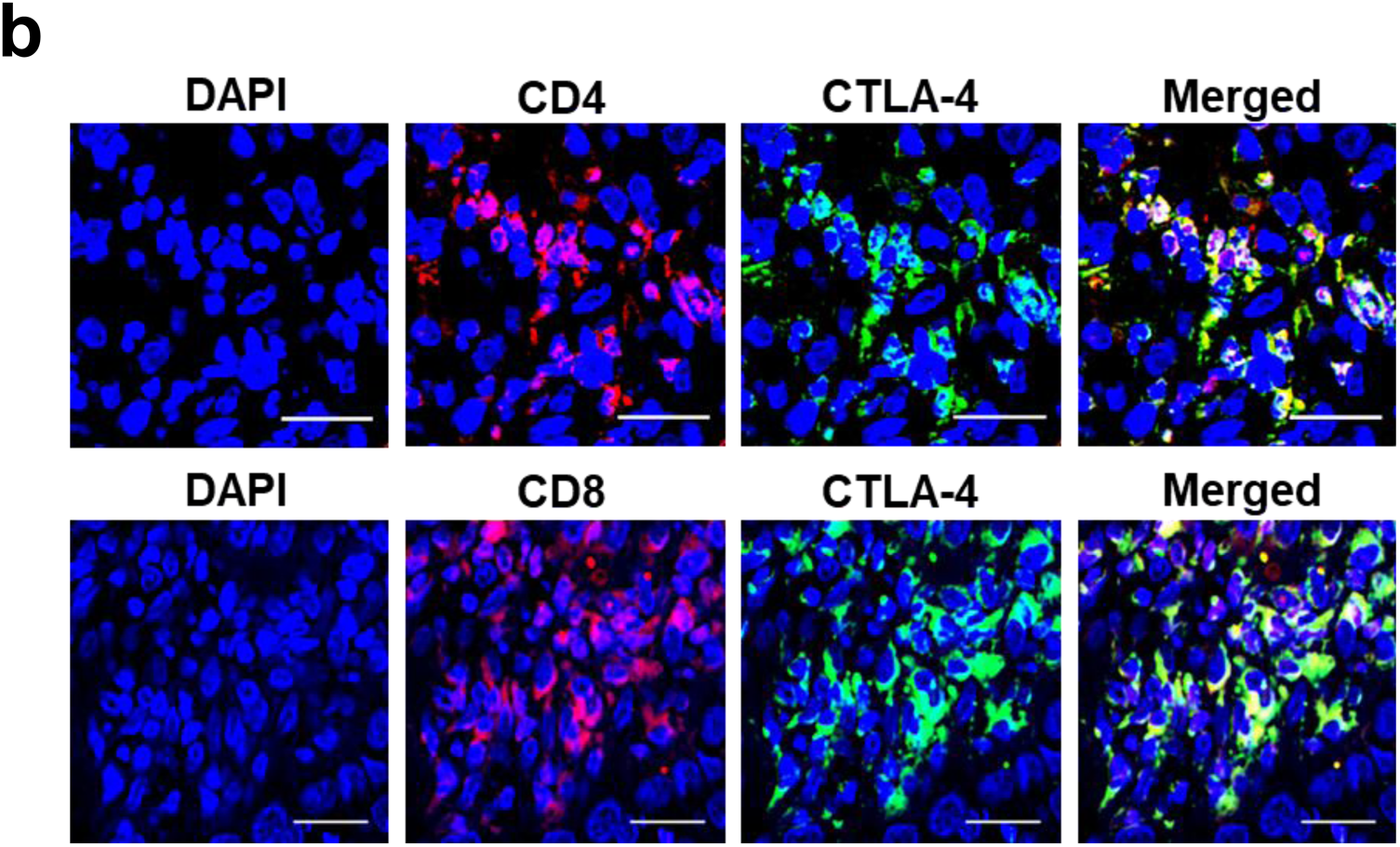
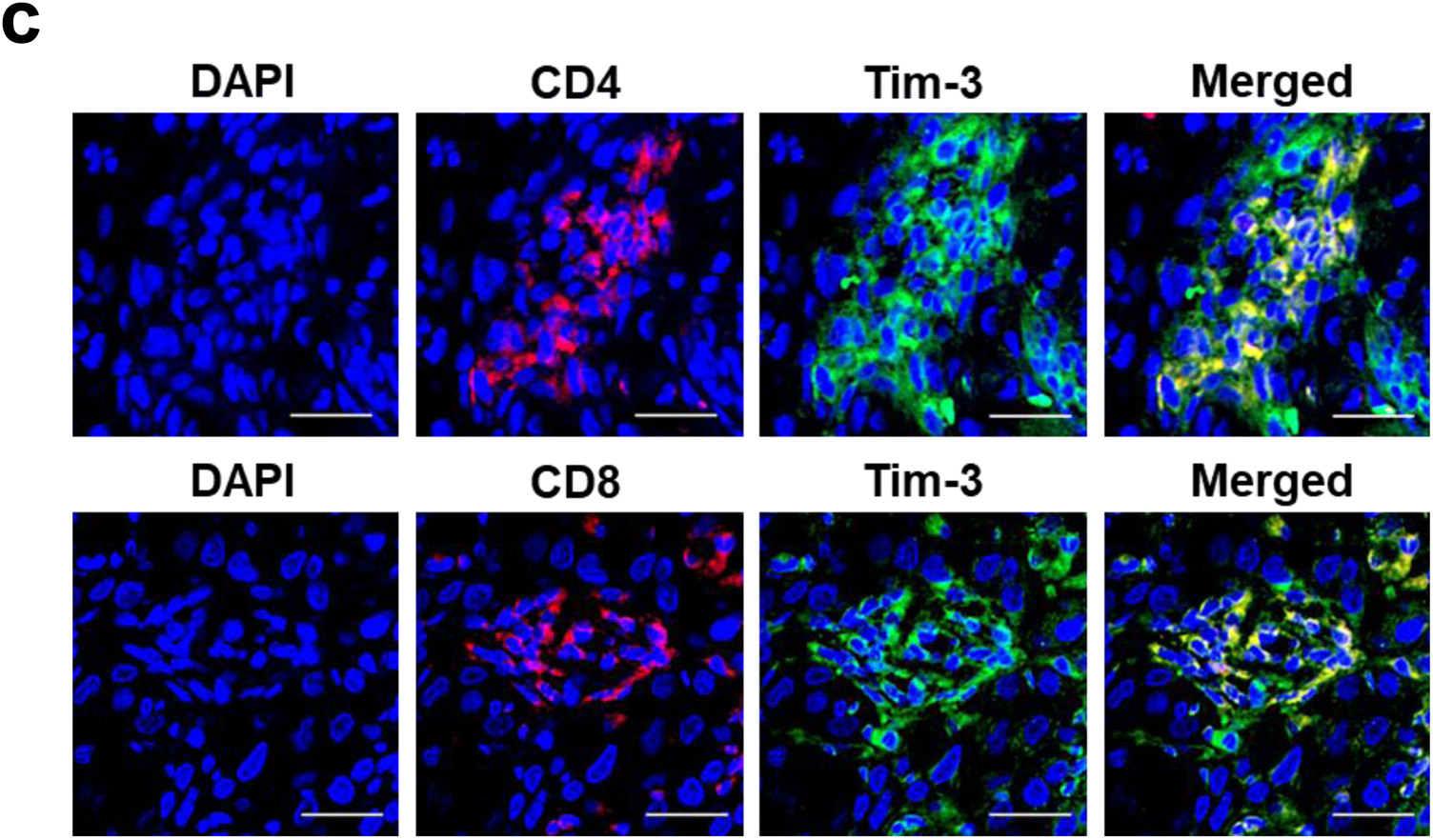
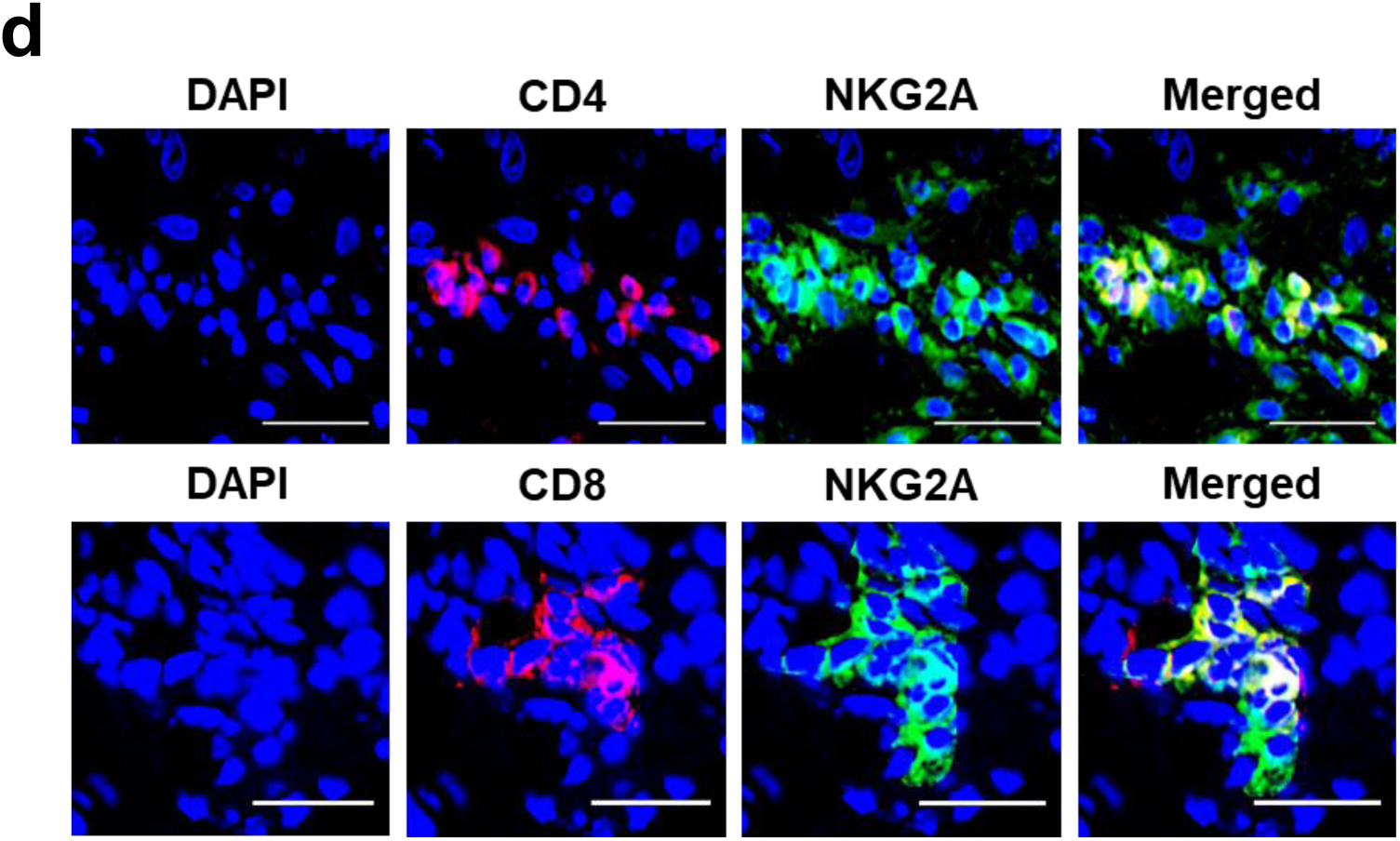
Representative images showing expression of various immune checkpoint proteins by CD4 and CD8 lymphocytes. FFPE sections of OSCC tumour tissues were simultaneously immune-stained with antibodies against various immune-checkpoint proteins and those against CD4 and CD8 lymphocytes (scale bar 10µm).

