## Supplementary Table 1 for "A pro-oxidant combination of resveratrol and copper down-regulates hallmarks of cancer and immune checkpoints in patients with advanced oral cancer: Results of an exploratory study (RESCU 004)"

Supplementary Table 1: Treatment regime

| **Doses Levels** | **Resveratrol** | **Copper** | **Patients (no.)** |
| --- | --- | --- | --- |
| Control | 0 | 0 | 5 |
| Dose level I | 5.6 mg | 560 ng | 5 |
| Dose level II | 50 mg | 5 µg | 5 |
| Dose level III | 500 mg | 50 µg | 5 |
| Dose level IV | 500 mg | 5 mg | 5 |
