## Supplementary Table 2 for "A pro-oxidant combination of resveratrol and copper down-regulates hallmarks of cancer and immune checkpoints in patients with advanced oral cancer: Results of an exploratory study (RESCU 004)"

**Supplementary Table 2:** Cancer hallmarks and corresponding biomarkers

| **Hallmark** | **Bio-marker** |
| --- | --- |
| **Sustained Proliferation** | TGF-β  c-Myc  pAKT |
| **Evading growth suppressor** | pATM  p53 |
| **Tumor promoting inflammation** | TNF-α  IL-6  NF-κβ  IFN-γ |
| **Enabling replicative immortality** | Cyclin D1 |
| **Avoiding immune destruction** | PD-1  PD-L1  CTLA-4  NKG2A  TIM-3 |
| **Inducing angiogenesis** | VEGFA |
| **Deregulating cellular energetics** | GLUT1 |
| **Resisting Cell Death** | Bcl-2 |
| **Genome instability and mutation** | Rad50  γH2AX |
| **Activating invasion and metastasis** | Vimentin  N-cadherine  MMP13 |
