## Supplementary Table 3.a, 3.b, 3.c for "A pro-oxidant combination of resveratrol and copper down-regulates hallmarks of cancer and immune checkpoints in patients with advanced oral cancer: Results of an exploratory study (RESCU 004)"

**Supplementary Table 3a:**

**Primary antibodies for antioxidant enzymes**

| **Sr. No** | **Antibody** | **Source** | **Catalogue No.** |
| --- | --- | --- | --- |
| 1 | Anti-SOD | ab13498 | Abcam, Cambridge, UK |
| 2 | Anti-Catalase | ab16731 | Abcam, Cambridge, UK |
| 3 | Anti-GPx | sc-133160 | Santa-Cruz Biotechnology, USA |

**Supplementary Table 3b:**

**Primary antibodies for cancer hallmark biomarkers**

| **Hallmark** | **Biomarker** | **Catalogue No.** | **Company / vendor** |
| --- | --- | --- | --- |
| **Sustained Proliferation** | TGF-β | T0438 | Sigma-Aldrich, USA |
|  | c-Myc | ab32072 | Abcam, UK |
|  | p-Akt | 9271S | Cell Signalling Technologies, USA |
| **Evading Growth Suppressors** | pp53 | sc-18078 | Santa Cruz Biotechnology, USA |
|  | pATM | 05-740 | Merck-Millipore, Germany |
| **Avoiding Immune Destruction** | PDL-1 | PRS-4059 | Sigma-Aldrich, USA |
|  | PD-1 | ab52587 | Abcam, UK |
|  | CTLA-4 | NBP2-42630 | Novus Biologicals, USA |
|  | NKG2A | PA5-21949 | Thermo-Scientific, USA |
|  | Tim-3 | ab47997 | Abcam, UK |
| **Enabling Replicative Immortality** | CCND-1 | sc-246 | Santa Cruz Biotechnology, USA |
| **Tumour Promoting Inflammation** | TNF-α | ab9739 | Abcam, UK |
|  | NF-кB | ab32536 | Abcam, UK |
|  | IL-6 | NB6001131 | Novus Biologicals, USA |
|  | IFN-**γ** | PA1-24782 | Thermo- Scientific, USA |
| **Invasion and Metastasis** | MMP13 | sc-12363 | Santa Cruz Biotechnology, USA |
|  | Vimentin | 5741 | Cell Signalling Technologies, USA |
|  | N-cadherin | 13116S | Cell Signalling Technologies, USA |
| **Inducing Angiogenesis** | VEGFA | ab 52917 | Abcam, UK |
| **Genome Instability and Mutation** | γH2AX | 05-636 | Merck-Millipore, Germany |
|  | Rad50 | sc-20155 | Santa Cruz Biotechnology, USA |
| **Resisting Cell Death** | Bcl-2 | s-492 | Santa Cruz Biotechnology, USA |
| **Deregulating Cellular Energetics** | Glut-1 | ab652 | Abcam, UK |

**Supplementary Table 3c:**

**Secondary Antibodies:**

| **Sr. No** | **Antibody** | **Source** | **Catalogue No.** |
| --- | --- | --- | --- |
| 1 | Goat Anti-Mouse IgG H&L (FITC) secondary Antibody | ab7064 | Abcam, Cambridge, UK |
| 2 | Rabbit Anti-Goat IgG H&L (FITC) secondary Antibody | ab6737 | Abcam, Cambridge, UK |
| 3 | Goat anti-Mouse IgG H& L (FITC) secondary Antibody | ab6785 | Abcam, Cambridge, UK |
| 4 | Goat anti-Rabbit IgG H&L (TRITC) secondary Antibody | ab6718 | Abcam, Cambridge, UK |
| 5. | Goat anti-mouse IgG H&L (TRITC) | ab6786 | Abcam, Cambridge, UK |
